# Intracranial Plaque Characteristics of Recurrent Ischemic Stroke After Intensive Medical Therapy for a 6-month Follow-up

**DOI:** 10.1101/2023.12.20.23300341

**Authors:** Zhang Shi, Boyu Zhang, Ming Zhao, Shujie Zhang, Jing Li, Qi Liu, Mengsu Zeng, Jiang Lin, Jianping Lu, He Wang

## Abstract

**BACKGROUND:** Intensive medical management has been recommended to ischemic stroke of intracranial atherosclerosis (ICAS), but 9.4-15% probability of recurrent stroke remains an inevitable reality. The characteristics of high-risk intracranial plaque that contribute to stroke recurrence after intensive therapy is unclear.

**METHODS:** All the patients for acute ischemic stroke due to ICAS underwent the 3D head&neck high-resolution vessel wall magnetic resonance imaging (hr-VW-MRI) at baseline and received intensive medical management within 90 days in this two-center and prospective study. The clinical risk factors and blood biomarkers were recorded. The morphological features, such as minimal lumen area (MLA), and histogram parameters including entropy were assessed based on MR images. The primary endpoint event after 6 months is recurrence of ischemic events (hyperintense signals on diffusion-weighted images or TIA in the ipsilateral vascular territory). Cox regression analysis was used to calculate the hazard ratio (HR) and 95% confidence interval (CI) for recurrent events.

**RESULTS:** 222 patients (age, 59.5±12.1; males, 153) were included in the subsequent analysis. After an average of six months of follow-up (average days, 177.4±23.1), 38 patients have reached the primary endpoint. After adjusting the age, BMI and gender, the multivariate Cox regression demonstrated that three clinical factors including smoking (HR=4.321; 95%CI, 1.838-10.161; *P*=0.001), taking exercise (HR=0.409; 95%CI, 0.198-0.843; *P*=0.015) and blood pressure management (HR=0.180; 95%CI, 0.073-0.443; *P*=0.001), and two MR-related parameters (MLA [HR=0.771; 95%CI, 0.625-0.951; *P*=0.015] and entropy [HR=0.274; 95%CI, 0.130-0.576; *P*=0.001]) were significant predictors of recurrent ischemic stroke. The Kaplan– Meier curve depicted that the cumulative incidences of patients with all high-risk features were significantly higher than those without(P < 0.001).

**CONCLUSIONS:** The plaque characteristics based on 3D head&neck hr-VW-MRI may provide complementary values over traditional clinical features in predicting ischemic recurrence for ICAS and help risk stratification of patients at risk of recurrent stroke.

## INTRODUCTION

Intracranial atherosclerosis (ICAS) has been recognized as the most common cause of ischemic stroke worldwide.^1^ Despite intensive management of modifiable risk factors and dual antiplatelet therapy has been recommended after ischemic stroke of atherosclerotic origin,^2,3^ the recurrence of ischemic stroke remains an inevitable reality. Following intensive lipid-lowering therapy, patients exhibit a 9.4% probability of experiencing the recurrent ischemic cerebrovascular event in the short term, and a 15.1% recurrence rate after one year, concurrently maintaining a high incidence of neurological disability.^4–6^ It is suggested that beyond managing the high-risk characteristics like blood lipid level, additional factors contribute to the recurrence of ischemic stroke, such as features of atherosclerotic plaque, remain to be elucidated.

High-resolution vessel wall magnetic resonance imaging (hr-VW-MRI) possesses the capability to identity plaque morphology and various atherosclerotic components of carotid and intracranial arteries, such as vessel wall remodeling, intraplaque hemorrhage, and plaque enhancement when a contrast agent is administered.^7,8^ These features have been used to estimate the plaque vulnerability and exhibit a significant association with future ischemic stroke events.^9^ Nevertheless, the relationship of the features of atherosclerotic plaque, particularly from intracranial artery, with recurrence of stroke had less been investigated by most of randomized controlled trials (RCTs) and previous cross sectional studies because of the imaging protocols from computed tomography angiography (CTA) or MR angiography (MRA).^4,10,11^ Additionally, some recent studies investigating changes in plaque features over time and their relationship to recurrent stroke were limited as follows: 1) small sample size ^12,13^; 2) the predominant use of two-dimensional (2D) MRI sequences, which have very limited spatial resolution in the slice-select direction, long acquisition times, and the inability to accurately portrait vessels along various orientations due to fixed acquisition orientation ^14^; and 3) most focused on lesion morphological and compositional features, lacking detailed descriptions of changes in plaque signal intensity and intra-structure^15^.

In this two-center, prospective study, based on advances in three-dimensional (3D) MR technology^16^ and our previous study on histogram of ICAS^17^, multivariate analysis including lifestyle habits, clinical features, and 3D hr-VW-MRI parameters including morphological and histogram characteristics were utilized to explore the complex nature of stroke recurrence and identify the high-risk factors associated with the recurrence of ischemic events.

## METHODS

### Data Availability

Anonymized data not published within this article can be provided upon request by the corresponding author.

### Participants

This study was approved by the Institutional Review Board of Changhai Hospital of Shanghai (No.CHEC2018-092) and Zhongshan Hospital, Fudan University (No.B2022-112R), and written informed consent was obtained from all patients. This prospective study was an observational cohort study^18^, which included patients admitting to Changhai Hospital of Shanghai from January 2019 to March 2021 and Zhongshan hospital from March 2022 to January 2023 for acute ischemic stroke due to ICAS. And all the patients underwent a 3D head&neck hr-VW-MRI prior to admission and had intensive medical management within 90 days (dual antiplatelet therapy [clopidogrel at an initial dose of 300 mg, followed by 75 mg/d for 90 days, plus aspirin at a dosage of 100 mg/d] and high-intensity statins [40 mg daily]) according to previous studies^19,20^ and guidelines^21,22^.

The inclusion criteria were as follows: 1) acute ischemic stroke (having acute ischemic symptoms and diffusion weighted imaging [DWI] positive for no more than 2 weeks), 2) intracranial arterial focal wall thickness >1mm on hr-VW-MRI^23^, and 3) more than one atherosclerotic risk factor, including hypertension, diabetes mellitus, hypercholesterolemia, or cigarette smoking.

Patients with the following conditions were excluded: 1) subacute stroke or transient ischemic attack (TIA); 2) underwent head or neck arterial stenting after a period of in-hospital intensive treatment; 3) hr-VW-MRI revealed significant stenosis of the extracranial carotid arteries (stenosis ≥50%) and/or the presence of vulnerable carotid plaques including a lipid-rich necrotic core on T1-weighted fat-suppressed images, intraplaque hemorrhage (IPH) and obvious enhancement;^24^ 4) presence of ascending aortic arch atheroma as identified on MRA (defined as plaque thickness > 4 mm);^25^ 5) nonatherosclerotic intracranial arterial disease including aneurysms with intervention therapy, vasculitis, moyamoya disease, dissection, reversible cerebral vasoconstriction syndrome, and intracranial dolichoectasia; 6) suspected cardiogenic thrombosis as assessed on cardiac Doppler ultrasound or cardiac CTA; 7) known coagulopathy; 8) heart failure or respiratory failure; 9) renal dysfunction (serum creatinine >133 μmol/liter); 10) serious disturbance of consciousness; 11) intracranial hemorrhage; and 12) clinical contraindications to MRI, such as patients with pacemakers, certain types of metallic implants, or severe claustrophobia.

### Clinical Data Collection

The demographic and clinical characteristics, including age, sex, body mass index, hypertension, hyperlipidemia, diabetes mellitus, smoking, and medication usage, were collected from the clinical record. According to these clinical features, ABCD^2^ scale were be calculated.^26^ All patients underwent NIHSS, Glasgow Coma Scale, and Modified Rankin Scale assessment upon admission and post-treatment at follow-up.^27^

Lab examinations of blood samples, which are highly related to atherosclerosis, were collected at admission as follows: fasting blood glucose, glycated hemoglobin, hemoglobin, cholesterol, triglycerides, low-density lipoprotein, high-density lipoprotein, apolipoprotein A1, apolipoprotein B, homocysteine, serum uric acid, erythrocyte sedimentation rate, lipoprotein(a), and C-reactive protein.

### Follow-up Data Collection

1. Evaluation of exercise: Post-discharge physical activities were assessed referring to the 2015 standards by the American College of Sports Medicine.^28^ Those who consistently engaged in physical activities more than 3 days a week were considered to take exercise. By inquiring, it was documented.
2. High-risk control: Patients with hypertension and diabetes were further grouped according to the following management types before admission: 1) no management, if the patient did not receive any treatment or had irregular treatment/monitoring; and 2) strict control, when the patient received treatment and monitoring regularly for more than 5 days per week.
3. Definition of ischemic recurrence: Recurrent ischemic events referred to the incidence of subsequent ischemic stroke (evidenced by hyperintense signals on DWI images) or TIA in the ipsilateral responsible vascular territory. Such recurrence of ischemic stroke/TIA was designated as the endpoint event for the current research. The assessment and collection of follow-up information were conducted by a neurologist by clinical visit or telephone interview, who was blind to the patients’ MRI results.

### MRI Acquisitions

Patients in Changhai Hospital underwent MRI scan on 3.0-T whole-body MR systems (MAGNETOM Skyra, Siemens Healthcare, Erlangen, Germany) using a 20-channel head coil combined with an 8-channel neck coil. And the MRI data from Zhongshan Hospital were acquired on a 3.0-T MRI scanner (uMR 790, United Imaging Healthcare, Shanghai, China) with a 24-channel receiver head coil combined with an 8-channel neck coil. Both acquisitions were kept for the head and neck joint scanning. The MRI protocol included T1-weighted imaging, post-contrast T1-weighted imaging, fluid-attenuated inversion recovery (FLAIR), DWI and time-of-flight MRA with more protocol details in the Table-S1. Post-contrast T1-weighted images were acquired using a dose of 0.2 mmol/kg Gd-DTPA injected by power injector at a rate of 2 mL/seconds followed by 15 mL of physiological saline.

### Imaging Analysis

All the MR images were analyzed by two experienced radiologists in vessel wall imaging (one with 8 years’ experience, and one with 12 years’ experience). The culprit vessel was defined as the artery arising on the ipsilateral side to a fresh infarction on the DWI images with accompanying clinical symptoms. Any disagreement was resolved by consensus.

The VesselMass software (Leiden University Medical Center, The Netherlands) was employed for semi-automatic delineation of plaques and automatic alignment of hr-VW-MRI scans encompassing T1-weighted imaging, post-contrast T1-weighted imaging and T2-weighted imaging. Manual correction was performed on all thickened vessel wall slices to obtain the inner and outer boundary at the proximal and distal non-stenotic slices of the lesion. Various metrics related to the plaque were calculated, primarily comprising quantitative metrics such as, minimum luminal area (MLA) stenosis rate (SR), plaque volume (PV), enhancement ratio (ER), plaque burden (PB), remodeling ratio (RR), eccentricity index (EI), and qualitative metrics including intraplaque hemorrhage (IPH). (The definitions of these features shown in Supplementary Files)

The histogram features of plaques were quantified using methods from our previous study.^17^ The mean, standard deviation (SD), minimum, maximum, medians, coefficient of variation (CV) and entropy values of each plaque region were calculated.

### Reproducibility

To evaluate the reproducibility of the imaging measurements, inter-reader agreement in measuring quantitative plaque morphology was evaluated on baseline images selected at random from 60 patients in the study population. And images were reviewed by one reader with one with 8 years’ experience twice with an interval of 12 weeks to avoid bias. The same MR images were independently reviewed by another reader with 12 years’ experience for testing the inter-observer agreement during the first time measured by the first reader.

### Statistical Analysis

1. The calculation of sample size: it was based on a two-sample unpaired t-test with 0.80 power and 0.05 significance level (two sided). The measurement error was used as the coefficient of variation for sample size determination. According to a previous scan-rescan study of intracranial vessel wall imaging,^29^ the coefficients of variation of measurements are 5%-10%. Hence 7.5% was used as the estimated coefficient of variation to detect 15% difference in the recurrent stroke group compared to stable group. The required sample size was eight in each group. Considering the recurrent stroke rate is 20% in our study cohort in 4 years, a sample size of 40 patients is needed (with eight recurrent strokes in 4 years).
2. All statistical analysis was undertaken using SPSS (version 24.0, IBM, Chicago, IL). The mean and SD were recorded for continuous variables, and the frequency and percentage were recorded for categorical variables. Univariable analysis was first performed. Two-sample t-test, chi-square test or Mann-Whitney U-test was applied to evaluate group differences according to requirement. A Kaplan-Meier survival analysis was performed to estimate the cumulative event-free rates. Univariable and multivariable Cox regression analyses were used to calculate the hazard ratio (HR) and corresponding 95% confidence interval (CI) of the plaque features in discriminating between patients with and without recurrent events. Variables with P < 0.10 in univariable analysis were selected as inputs for the multivariable model and then underwent sequential backward elimination to a P value < 0.05. The diagnostic performance was described using receiver operating characteristic (ROC) curves and area under curve (AUC) values. The inter-reader reproducibility of continuous variables was evaluated using the intra-class coefficient (ICC) with a two-way random-effects model with absolute measurements, while the Kappa value was determined for the categorical variables. All tests were two-sided and a P value < 0.05 was considered significant.

## RESULTS

The patient characteristics are presented in Table 1 and Figure-2 describes the workflow of the inclusion and exclusion process of the patients. A total of 250 patients were initially recruited, and 28 patients were excluded due to ≥70% stenosis of the extracranial carotid arteries (n=6), presence of vulnerable carotid plaque (IPH) but with ≥50% stenosis of the extracranial carotid arteries (n=5), intracranial dissection (n=5), imaging quality score 2 (n=5), undergoing head or neck arterial stenting during in-hospital treatment (n=4), and patients with severe claustrophobia (n=3). Therefore, 222 patients (age, 59.5±12.1; males, 153) were included in the subsequent analysis. After 177.4 days (±23.1) of follow-up, 38 patients reached the primary endpoint.

**Figure-1:**
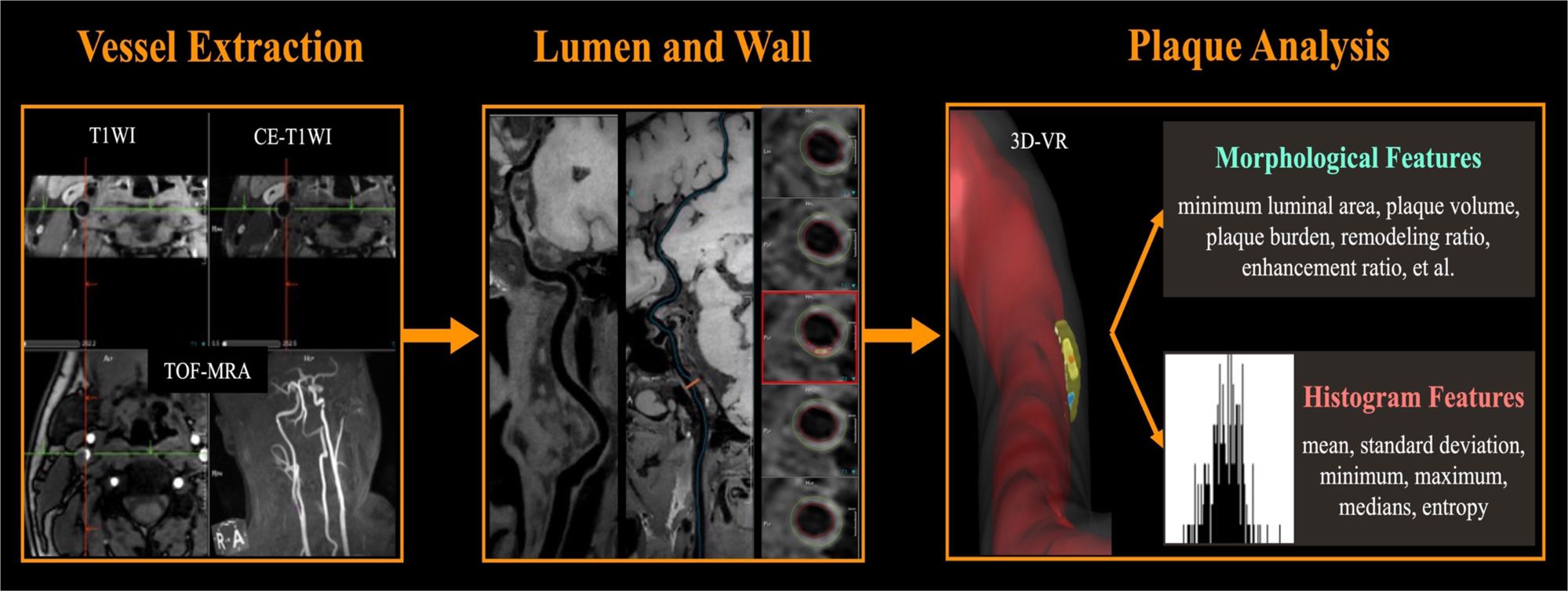
The procedure of the imaging analysis on high-resolution vessel wall magnetic resonance imaging.

**Figure-2:**
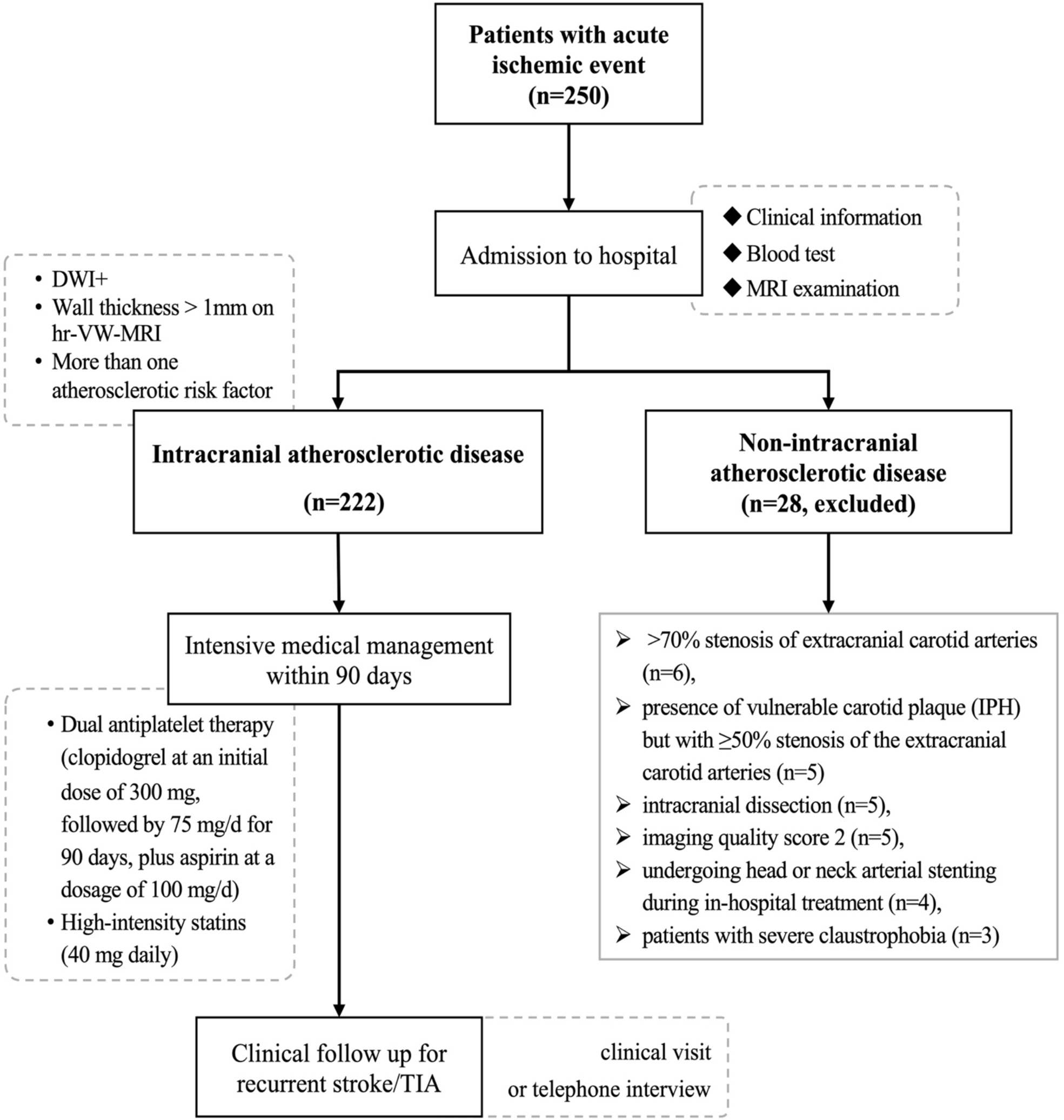
The flowchart of the study with patients’ recruiting criteria.

**Table1.** Demographic and clinical features associated with ischemic recurrence.

| Characteristics | Mean ± SD, median (IQR), or n (%) |  |  | P Value |
| --- | --- | --- | --- | --- |
|  | Total (n=222) | Non-recurrent (n=184) | Recurrent (n=38) |  |
| Age | 59.50±12.11 | 59.27±12.45 | 60.63±10.42 | 0.365 |
| Sex (Male) | 153 (68.9) | 117 (63.6) | 36 (97.4) | **<0.001** |
| BMI | 25.55±2.87 | 25.67±2.98 | 24.98±2.13 | 0.188 |
| Diabetes mellitus | 122 (54.9) | 96 (52.2) | 26 (68.4) | 0.067 |
| Hypertension | 170 (76.6) | 140 (76.1) | 30 (78.9) | 0.705 |
| Hyperlipidemia | 76 (34.2) | 64 (34.8) | 12 (31.6) | 0.848 |
| Smoking | 102 (45.9) | 78 (42.4) | 24 (63.2) | **0.019** |
| Drinking | 43 (19.4) | 31 (16.8) | 12 (31.6) | **0.036** |
| Taking exercise | 157 (70.7) | 141 (76.6) | 16 (42.1) | **<0.001** |
| Scores on admission |  |  |  |  |
| ABCD2 | 4.0 (3.0, 6.0) | 3.0 (2.0, 4.0) | 4.0 (3.0, 6.0) | **0.035** |
| NIHSS | 3.0 (1.0, 5.0) | 2.0 (0, 3.0) | 3.0 (2.0, 5.0) | **0.023** |
| GCS | 15.0 (15.0, 15.0) | 15.0 (15.0, 15.0) | 15.0 (15.0, 15.0) | 0.253 |
| ASPECTS | 10.0 (9.0, 10.0) | 10.0 (9.0, 10.0) | 10.0 (8.0, 10.0) | **0.032** |
| Lab examinations |  |  |  |  |
| Fasting glucose | 6.4 (5.3, 8.7) | 6.2 (5.3, 8.7) | 6.9 (5.3, 10.0) | 0.239 |
| Glycosylated hemoglobin | 6.2 (5.4, 9.2) | 6.2 (5.3, 9.0) | 9.2 (5.5, 9.7) | 0.101 |
| Hemoglobin | 141.0 (130.0, 152.0) | 142.0 (128.0, 153.0) | 139.0 (132.0, 150.0) | 0.885 |
| Total cholesterol, mmol/L | 4.53 (3.42, 5.56) | 4.58 (3.54, 5.58) | 4.10 (3.35, 5.50) | 0.217 |
| Triglyceride, mmol/L | 1.34 (1.19, 2.08) | 1.30 (1.07, 1.33) | 1.40 (1.20, 2.11) | **0.012** |
| LDL cholesterol, mmol/L | 2.94 (1.97, 3.70) | 2.99 (1.98, 3.70) | 2.51 (1.97, 3.99) | 0.505 |
| HDL cholesterol, mmol/L | 1.15 (1.03, 1.25) | 1.15 (1.02, 1.25) | 1.16 (1.06, 1.26) | 0.883 |
| ApoA1 | 1.10±0.16 | 1.09±0.15 | 1.12±0.17 | 0.480 |
| ApoB | 0.83±0.29 | 0.83±0.21 | 0.82±0.31 | 0.800 |
| Homocysteine | 10.25 (8.90, 11.45) | 10.15 (8.70, 11.60) | 10.09 (9.10, 11.30) | 0.049 |
| Uric Acid | 0.32 (0.26, 0.38) | 0.32 (0.26, 0.41) | 0.30 (0.22, 0.34) | **0.042** |
| ESR | 5.0 (3.0, 10.25) | 5.0 (3.0, 10.0) | 10.0 (5.2, 11.3) | **0.013** |
| Lipoprotein A | 16.6 (9.0, 38.08) | 16.4 (9.0, 37.2) | 20.5 (9.0, 91.6) | 0.123 |
| CRP | 3.2 (2.2, 5.4) | 3.3 (2.2, 5.6) | 3.2 (1.1, 3.8) | 0.145 |
| Blood pressure control | 164 (73.9) | 142 (77.2) | 22 (57.9) | **0.014** |
| Glucose control | 116 (52.3) | 92 (50.0) | 24 (63.2) | 0.139 |

BMI=body mass index; ABCD2=The model combining age, blood pressure, clinical features, diabetes and duration of systems; NIHSS=The National Institutes of Health Stroke Scale; GCS=Glasgow Coma Scale; ASPCETS=Alberta Stroke Program Early CT Score ; LDL=low-density lipoprotein; HDL=high-density lipoprotein; ESR=erythrocyte sedimentation rate; CRP=C-reactive protein

### Clinical characteristics and lifestyles associated with recurrent ischemic stroke

Table 1 showed the comparison of clinical characteristics between recurrent and non-recurrent patients. Compared to non-recurrent patients, those with recurrent stroke had a larger proportion of males (63.6% vs 97.4%, *P*<0.001), smokers (42.4% vs 63.2%, *P*=0.019), alcohol consumers (16.8% vs 31.6%, *P*=0.036), and better blood pressure control (77.2% vs 57.9%, *P*=0.014), as well as lower proportion of maintain regular exercise (76.6% vs 42.1%, *P*<0.001). Moreover, the clinical scores during the hospital stay, including ABCD^2^ (*P*=0.035), NIHSS (*P*=0.023) and ASPECTS (*P*=0.032), were associated with recurrent ischemic events. Furthermore, in laboratory examinations, it indicated that the recurrent patients had lower levels of triglyceride (1.30 [IQR, 1.07 to 1.33] vs 1.40 [IQR, 1.20 to 2.11], *P*=0.012) and ESR (5.0 [IQR, 3.0 to 10.0] vs 10.0 [IQR, 5.2 to 11.3], *P*=0.013), but elevated levels of uric acid (0.32 [IQR, 0.26 to 0.41] vs 0.30 [IQR, 0.22 to 0.34], *P*=0.042).

### Morphological and histogram parameters based on MR predicting stroke recurrence

The MRI and corresponding histogram characteristics of the plaques are presented in Table 2. No significant differences were observed between patients with and without recurrence in terms of their culprit vessels. Regarding morphological features, MLA was the only predictor of the recurrence, and the findings showed the recurrent patients exhibited smaller MLA (3.48±0.23mm^2^ vs 2.44±0.14mm^2^, *P*=0.010) than those without ischemic recurrence. Histogram features based on T1W images, such as SD (*P*=0.005), maximum value (*P*=0.018), coefficient of variation (*P*=0.031) and entropy (*P*=0.002), were significantly associated with stroke recurrence.

**Table2.** Morphological and histogram characteristics associated with ischemic recurrence.

| Characteristics | Mean ± SD, median (IQR), or n (%) | | P Value |
| --- | --- | --- | --- |
|  | Non-recurrent<br>(n=184) | Recurrent (n=38) |  |
| Culprit vessels |  |  | 0.795 |
| ICA | 29 (15.8) | 6 (15.8) |  |
| MCA | 57 (31.0) | 12 (31.6) |  |
| ACA | 0 (0) | 4 (10.5) |  |
| BA | 86 (46.7) | 10 (26.3) |  |
| VA | 6 (3.3) | 4 (10.5) |  |
| PCA | 6 (3.3) | 2 (5.3) |  |
| Morphological features |  |  |  |
| Thickness wall (mm) | 1.15±0.32 | 1.09±0.26 | 0.304 |
| MLA (mm<sup>2</sup>) | 3.48±0.23 | 2.44±0.14 | **0.010** |
| Plaque volume (mm<sup>3</sup>) | 99.30 (59.23, 168.0) | 87.30 (50.0, 172.10) | 0.412 |
| Stenosis ratio (%) | 61.61±8.59 | 61.66±9.32 | 0.975 |
| Plaque burden (%) | 76.89±11.12 | 79.91±9.48 | 0.089 |
| Enhancement ratio (%) | 28.06 (9.44, 62.72) | 36.19 (13.98, 55.75) | 0.536 |
| Eccentricity ratio (%) | 60.24±20.05 | 64.43±15.16 | 0.224 |
| Remodeling index (%) | 97.41±31.53 | 100.64±31.59 | 0.568 |
| IPH | 29 (15.8) | 4 (10.5) | 0.409 |
| Histogram features |  |  |  |
| Means | 115.73±39.28 | 106.29±26.64 | 0.159 |
| Standard deviation | 28.03 (20.71, 37.27) | 21.53 (16.0, 27.51) | **0.005** |
| Minimum value | 55.58±4.74 | 48.41±2.24 | 0.364 |
| Maximum value | 108.89±5.65 | 157.84±4.19 | **0.018** |
| Medians | 155.24±39.75 | 106.47±28.33 | 0.198 |
| Coefficient of variation | 0.25 (0.19, 0.33) | 0.21 (0.19, 0.24) | **0.031** |
| Entropy | 6.46±0.49 | 6.15±0.54 | **0.002** |

ICA=internal carotid artery; MCA=middle cerebral artery; ACA=anterior cerebral artery; BA=basilar artery; VA=vertebral artery; PCA=posterior cerebral artery; MLA=minimal lumen area; IPH= intraplaque hemorrhage

### Univariate and multivariate Cox regression of recurrent stroke

The univariate Cox regression analysis indicated that patients maintaining regular exercise (*P*=0.015), having higher ASPECTS scores (*P*=0.004), elevated triglyceride (*P*=0.003) and uric acid levels (*P*=0.006), better blood pressure management (*P*=0.015), greater MLA (*P*=0.004), higher maximum value (*P*=0.016), and increased entropy (*P*=0.001) were less likely to experience a recurrence of ischemic events (Table-S2). Conversely, smokers (*P*=0.008) exhibited an increased risk of recurrence. After adjusting for age, BMI and gender, the multivariate Cox regression demonstrated that smoking (*P*=0.001), taking exercise (*P*=0.015), blood pressure management (*P*=0.001), MLA (*P*=0.015), and entropy (*P*=0.001) were significant predictors of recurrent ischemic stroke (Figure-3A).

**Figure-3:**
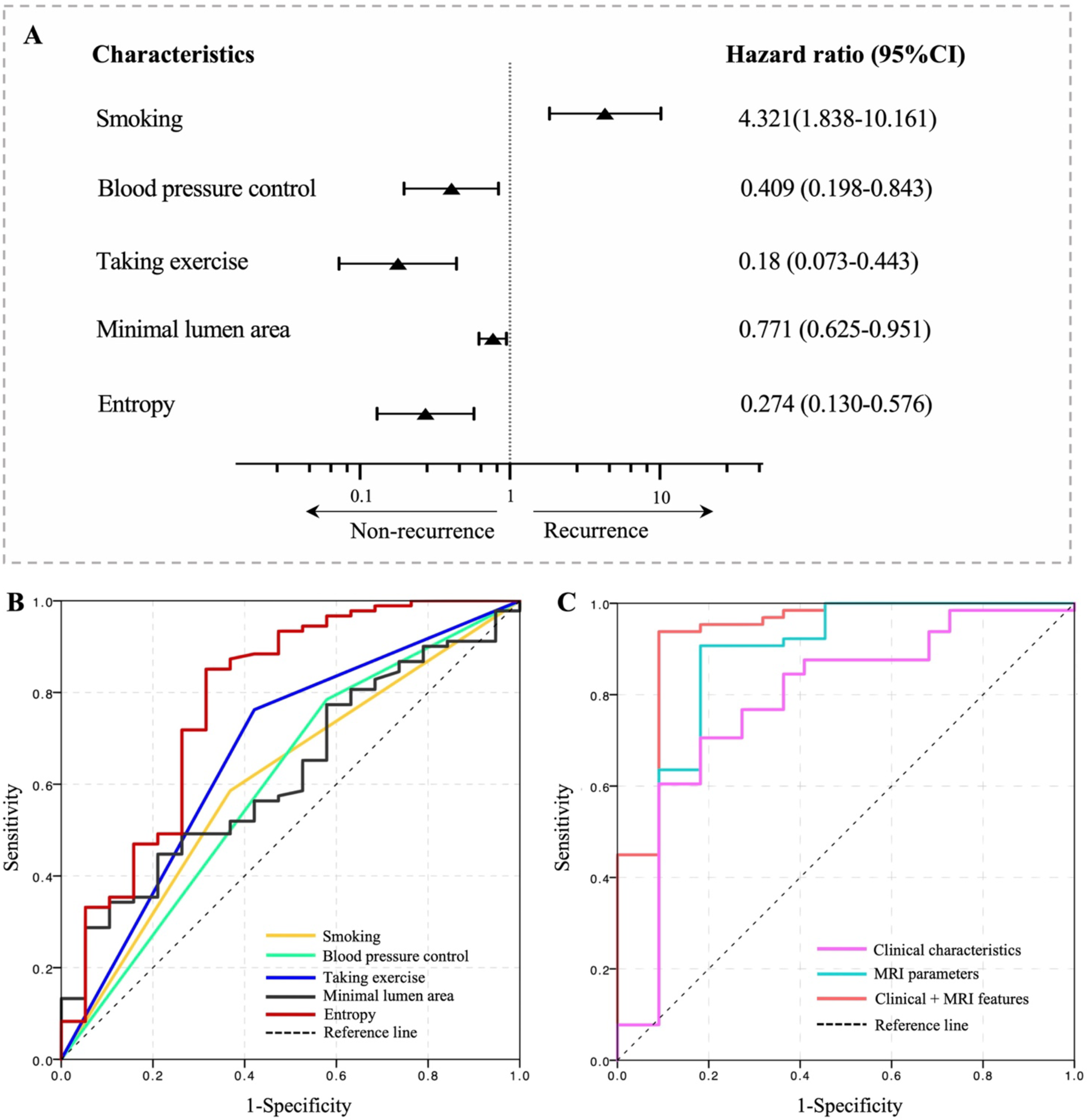
The forest plot and receiver operating characteristic curves associated with recurrent stroke for each parameter. (A) HR and 95% CI on the basis of multivariable Cox regression; (B) ROC curves for the risk factors identified by the univariate Cox regression analysis; and (C) ROC curves of combined models including traditional clinical features and MR-related parameters to predict recurrent TIA/stroke.

The AUC of ROC curves about the final Cox regression model, shown in Figure-3B, (including all three independent risk factors) was 0.885 (95% CI, 0.823-0.941), which was significantly higher than the AUC of any single factors (smoking: 0.609 [95%CI, 0.511-0.707], *P* =0.002; blood pressure control: 0.603 [95%CI, 0.499-0.707], *P* =0.001; taking exercise: 0.671 [95%CI, 0.571-0.770], *P* =0.013; MLA: 0.634 [95%CI, 0.584-0.713], *P* =0.025; entropy: 0.782 [95%CI, 0.690-0.875], *P* =0.038). Additionally, when comparing the AUC values, the traditional clinical factors had lower values than the MRI parameters (0.784[95%CI, 950.672-0.897] vs 0.859[95%CI, 0.745-0.973], *P* =0.01) as shown in Figure-3C.

The Kaplan–Meier curves of the five parameters for the recurrence of ischemic stroke were depicted in Figure-4A-E, and the cumulative incidences of patients with all high-risk features were significantly higher than those who had no high-risk features (*P* < 0.001; Figure-4F).

**Figure-4:**
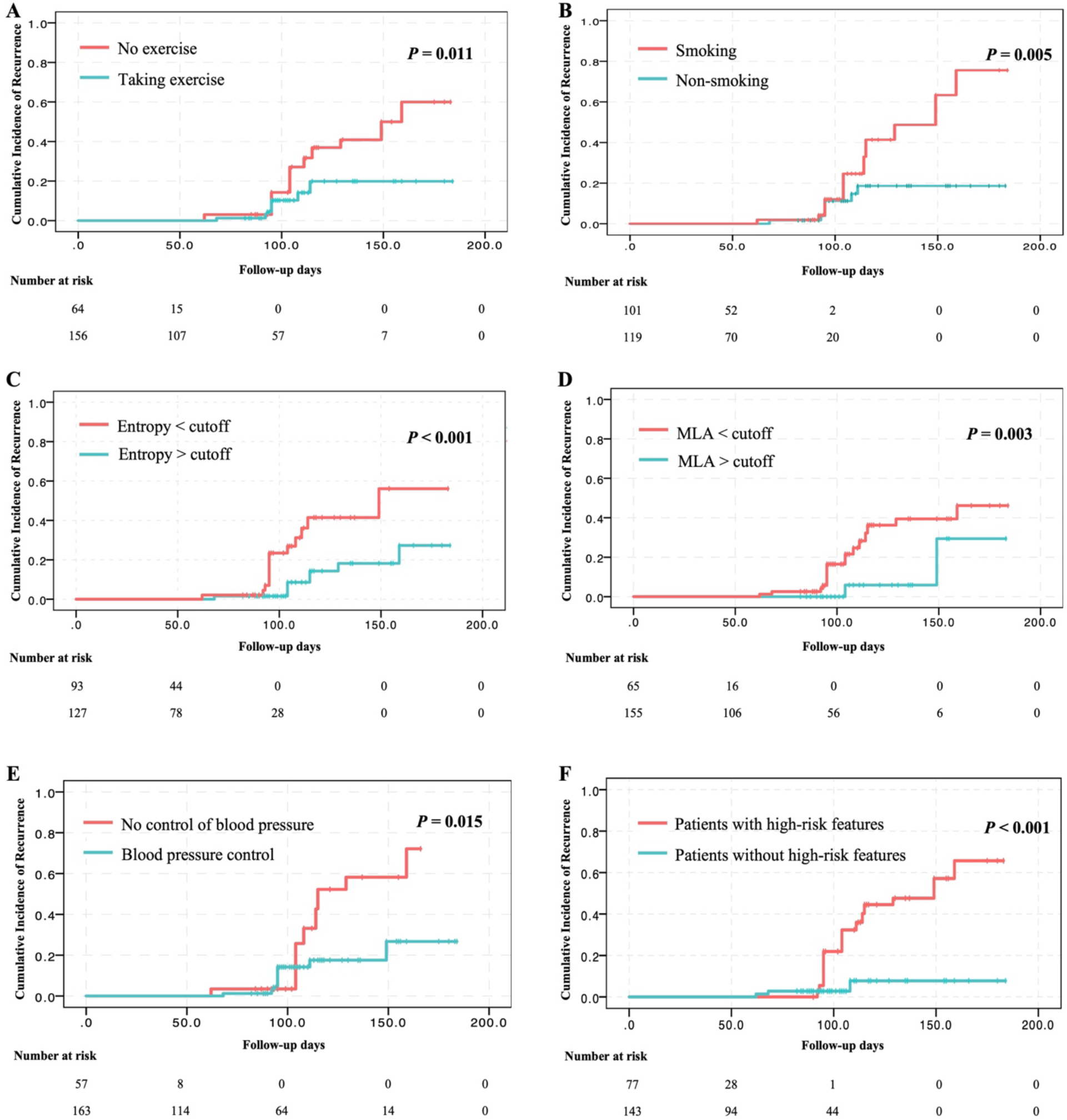
Kaplan–Meier curves of the five parameters (A-E) and the cumulative incidences of patients with all high-risk features (F) for the recurrence of ischemic stroke.

### Reproducibility

The reliability for the identification of morphological and compositional features in the plaque was assessed through the intra- and inter-observer agreement analysis, as presented in Supplementary table.

## DISCUSSION

In the current study, maintaining regular exercise and proper blood pressure management were identified to be associated with reduced risk of ischemic recurrence after intensive treatment, whereas smoking was found as a factor that elevated the recurrent risk. Moreover, this study indicated that the MLA and entropy of plaques were correlated with the recurrence of ischemic events. These findings provide preliminary data for managing recurrent ischemic events after intensive treatment from three aspects: lifestyle, clinical management, and plaque characteristics, thereby addressing potential factors influencing the recurrence of ischemic event recurrence after intensive medical treatment.

Lifestyle, encompassing aspects such as diet, physical activity, smoking, alcohol consumption, and sleep, are considered to be closely related to the recurrence of stroke.^30^ Consistent with previous studies, this study found that smoking amplified the risk of ischemic stroke recurrence. Previous research has identified that even 1 cigarette per day could significantly elevate the risk of stroke,^31^ and one genetic study involving large sample sizes has confirmed a potential causal relationship between smoking and stroke.^32^ In the Chinese population, a history of smoking has been correlated with a recurrence of stroke within 12 months.^33^ Moreover, previous study has suggested that smoking might increase the risk of stroke and its recurrence by inducing arterial stiffness and atherosclerosis.^34^ All of this evidence suggests the importance of managing cigarette use as a modifiable factor in the stroke recurrence. Furthermore, this study indicated that maintaining regular exercise contributed to reducing the risk of ischemic event recurrence. Previous studies have suggested that engaging in long-term, regular, moderate exercise is associated with a lower recurrence rate among stroke survivors, while irregular exercise is linked to an increased risk of stroke recurrence.^35^ In addition, moderate aerobic exercise has been found to improve cerebral perfusion and cognitive function in elder individuals.^36^ The beneficial effects of exercise on the brain might relate to several underlying mechanisms involving muscle–brain, liver–brain, and gut–brain crosstalk, which involve reducing neuroinflammation and enhancing neuroplasticity.^37^ Overall, maintaining exercise might be another modifiable lifestyle factor for stroke recurrence, but caution might be maintained in the choice of intensity and regularity of exercise.

Hypertension is one of the leading risks for stroke, making the control of high blood pressure a crucial intervention in secondary stroke prevention.^38^ Intensive blood pressure control has been found to reduce the risk of stroke recurrence and decelerate the progression of white matter lesions.^39^ Conversely, poor blood pressure control has been identified to be associated with elevated risk of first-ever stroke.^40^ Similarly, this study indicated that passive poor blood pressure control after intensive stroke treatment correlated with an increased risk of ischemic stroke recurrence. This result added to the evidence that favorable medication adherence might be one of the pivotal factors in preventing stroke recurrence. Besides, poor blood pressure control might also increase blood pressure variability, which has been suggested to exhibit adverse impact on the cerebrovascular system and increase the risk of stroke.^41,42^

In addition, this study utilized hr-VW-MRI to assess the associations of intracranial plaque morphologies with stroke recurrence and has demonstrated that a larger MLA was associated with a lower risk of recurrent ischemic events. In alignment with previous studies, a smaller lumen area of the middle cerebral artery might imply an increased risk of first-ever stroke.^43^ This result is much expected, as a larger MLA suggested a comparatively lesser degree of lumen constriction, thus potentially exerting a smaller impact on blood flow, mitigating the recurrence of ischemic events. For instance, one research has found that compared to patients with asymptomatic middle cerebral artery stenosis, those with symptomatic middle cerebral artery stenosis exhibit higher wall shear stress at the MLA.^44^ However, some existing studies had demonstrated other imaging features significantly associated with the recurrent ischemic stroke, such as plaque contrast enhancement,^15,41^ positive remodeling,^45^ plaque bureden,^14^ and the location of the intracranial plaques.^46^ The variability and mechanisms of stroke recurrence due to different plaque morphologic features still need to be revealed by more future studies.

In the present study, histogram features were performed to assess the change of the signal intensity on the intracranial atherosclerotic plaque, which could indirectly show the inter-structure of the plaque. Our previous studies of histogram features on intracranial plaque have better facilitated the differentiation of culprit and non-culprit plaques more than general morphological features. Furthermore, our findings in this study demonstrated that the entropy of plaques stood as an independent factor influencing the recurrence of ischemic events, with lower entropy suggesting an increased risk of recurrence. Entropy, which describes the uncertainty of a complexity system, is believed to reflect the level of details within the system to be observed.^47^ Under the present situation, the more complex and loose the structures of the constituent molecules, the larger the entropy of its histogram is. Although the exact mechanism is unclear, it may be related to atherosclerotic inclusions in the lesion. According to in-vivo studies with atherosclerotic plaques from the carotid artery and ex-vivo studies with lesions from MCA, hr-VW-MRI provides contrasts which could differentiate various atherosclerotic components in the lesion.^17^ Hence, lower entropy might imply a more homogeneous structure within the plaque, typically characterized by clustered foam cells, thereby rendering the plaque more active and making it greater risk of rupturing and causing a stroke.^48^

Despite the exciting findings, there are several limitations to be addressed. First, this study is limited to Southeast Chinese populations. As a result, findings presented in the current study might not pronounce in the national or global population and should also be interpreted with caution. Future studies involving multi-racial individuals are required to test the reliability of the current results. Secondly, there was a small sample size of recurrent stroke population in this study (n=38), and the time span of this study was nearly four years from 2019 to 2023, due to the COVID-19 pandemic. The examination of hr-VW-MRI was stopped during the most of patients infected by SARS-CoV-2 in Shanghai. Furthermore, although 3D sequences were utilized in this study, enabling the analysis of volumetric features of plaques, a higher image resolution would still greatly benefit the study. This might lead to a situation where larger lesions are imaged effectively, while smaller lesions, which might also cause infarction, cannot be captured by current MRI techniques. Thus, urgent development in this aspect is needed in further studies.

## ACKNOWLEDGMENT

None.

